# Why Patients Choose Spiritual Healers, Alternative Medicine, and Unqualified Practitioners Before Formal Medical Care: An Exploratory Mixed Methods Study in Peri-Urban and Rural Faisalabad, Pakistan

**DOI:** 10.64898/2026.04.23.26351601

**Authors:** Sadia Hamid, Muhammad Muneez, Shehzad Saleem

## Abstract

**Background:** Before obtaining professional medical care, many people in peri-urban and rural Pakistan contact herbalists, spiritual healers, and unlicensed caregivers. This study examined the social, economic, and cultural factors influencing the use of informal care by analysing the health-seeking behaviours of individuals in the Faisalabad District.

**Methods:** An exploratory mixed-methods study was conducted in Makkuana and the surrounding villages of Faisalabad District, Punjab. The quantitative component involved a cross-sectional survey of 69 adults using a structured questionnaire adapted from the I-CAM-Q. The qualitative component comprised twelve in-depth interviews and two focus group discussions. Descriptive statistics and chi-square analysis were used for quantitative data. Thematic analysis, guided by the Health Belief Model and Andersen’s Behavioural Model, was applied to qualitative data.

**Results:** The mean age of participants was 40.4 years; 62.3% were female, and 79.7% had monthly household incomes below PKR 60,000. Of the 69 participants, 68 (98.6%) sought care from an informal provider first, most commonly an unqualified practitioner (50.7%), herbal practitioner (29.0%), or homeopath (17.4%). Trust was the leading reason for provider choice (43.5%), followed by proximity (24.6%) and low cost (15.9%). Complications were reported by 21.7% of participants, and 39.1% later required formal care for the same illness. Eight qualitative themes emerged: structural and economic barriers to formal care; proximity and convenience as determinants of informal care; trust, familiarity, and social networks; cultural and religious normalisation of traditional practices; poor doctor-patient communication in formal settings; perceived safety and naturalness of alternative remedies; awareness deficits about provider qualifications; and treatment-related harm and delayed escalation to formal care.

**Conclusion:** Informal health care seeking is nearly universal in this community, driven by intersecting economic, structural, cultural, and interpersonal factors. Enhancing primary care affordability, accessibility, and the quality of provider-patient communication together with culturally sensitive health literacy programs, is essential to redirect care seeking toward qualified providers.

## BACKGROUND

Traditional, complementary, and integrative medicine health systems worldwide, and the remain common worldwide, specially in low-resource areas where formal healthcare availability is difficult. The World Health Organization has called for evidence on patterns of traditional healthcare use and its interaction with biomedical services [1]. Many people seek treatment from unqualified providers, homeopath, herbal practitioner, spiritual healers in Pakistan due to a complex interaction of cultural beliefs, access limitations, cost considerations, and community trust networks [2,3].

Previous studies have shown that relying on informal healthcare can result in treatment delays,financial issues and serious complications, specially in serious and chronic illnesses like cancer [4,5]. Similarly, findings from Karachi’s research on mental health treatment pathways reveal that traditional healers frequently serve as the initial point of contact, and patients go back and forth between spiritual and medical providers before receiving the proper management [6].Despite these results, there limited mixed-methods research exploring why people in periurban and rural Punjab rely on informal healthcare provider before seeking formal care.

Makkuana and nearby villages in Faisalabad District are examples of places where formal and informal healthcare coexist, providing a chance to comprehend the social, cultural, and economic elements influencing treatment selection. Hence, this study aimed to explore why residents sought care from spiritual healers, alternative practitioners, and unqualified providers before formal medical care, and identified the linked factors.

## METHODS

### Study design and setting

An exploratory mixed-methods study with a convergent parallel design was conducted in Makkuana and the surrounding peri-urban and rural villages of Faisalabad District, Punjab, Pakistan, during March and April 2026.

### Participants and sampling

Adult residents aged 18 years or above who had experienced illness in the preceding twelve months and had lived in the study area for at least six months were eligible. Individuals unable to communicate effectively or those with severe acute illness were excluded. The sample size was calculated using the single population proportion formula with an estimated CAM use prevalence of 51.7% from a national survey [7], a 90% confidence level, and a margin of error of 0.10, resulting in a minimum sample size of 68 participants. The final quantitative sample was 69 participants. For the qualitative component, twelve in-depth interviews and two focus group discussions (six participants each) were conducted until thematic saturation was reached. Purposive sampling was used throughout.

### Data collection

After obtaining written informed consent, participants completed a structured questionnaire adapted from the International Complementary and Alternative Medicine Questionnaire (I-CAM-Q) [8], capturing socio-demographic data, health-seeking behaviour over the preceding twelve months, reasons for provider choice, treatment outcomes, complications, cost, and attitudes toward formal and alternative care. In-depth interviews and focus group discussions were conducted in Urdu and Punjabi by a trained researcher using a semi-structured guide developed from the Health Belief Model [9] and Andersen’s Behavioural Model of Health Service Utilisation [10], covering illness beliefs, reasons for informal provider choice, perceived treatment effectiveness, experiences with formal services, and social and financial impact.

### Data analysis

Quantitative data were analysed using SPSS version 27. Descriptive statistics, including frequencies, percentages, means, and ranges were calculated. Chi-square tests were performed to examine associations between socio-demographic variables (sex, education, income) and provider choice, and between provider type and treatment outcomes. A p-value of less than 0.05 was considered statistically significant. Qualitative transcripts were analysed thematically with codes developed inductively from participant narratives and grouped into higher-order themes. Trustworthiness was enhanced through triangulation of qualitative and quantitative data and independent coding by two researchers.

### Ethics

Ethical approval was obtained from the Institutional Review Board of King Edward Medical University, Lahore. Written informed consent was obtained from all participants. Participation was voluntary, and confidentiality was maintained throughout.

## RESULTS

### Socio-demographic characteristics

Sixty-nine participants were enrolled with a mean age of 40.4 years (range 19 to 71). The majority were female (43, 62.3%) and currently married (53, 76.8%). More than half of the population had primary education or below (36, 52.2%), and 79.7% reported a monthly household income below PKR 60,000. Socio-demographic characteristics are summarised in Table 1.

**Table 1.**
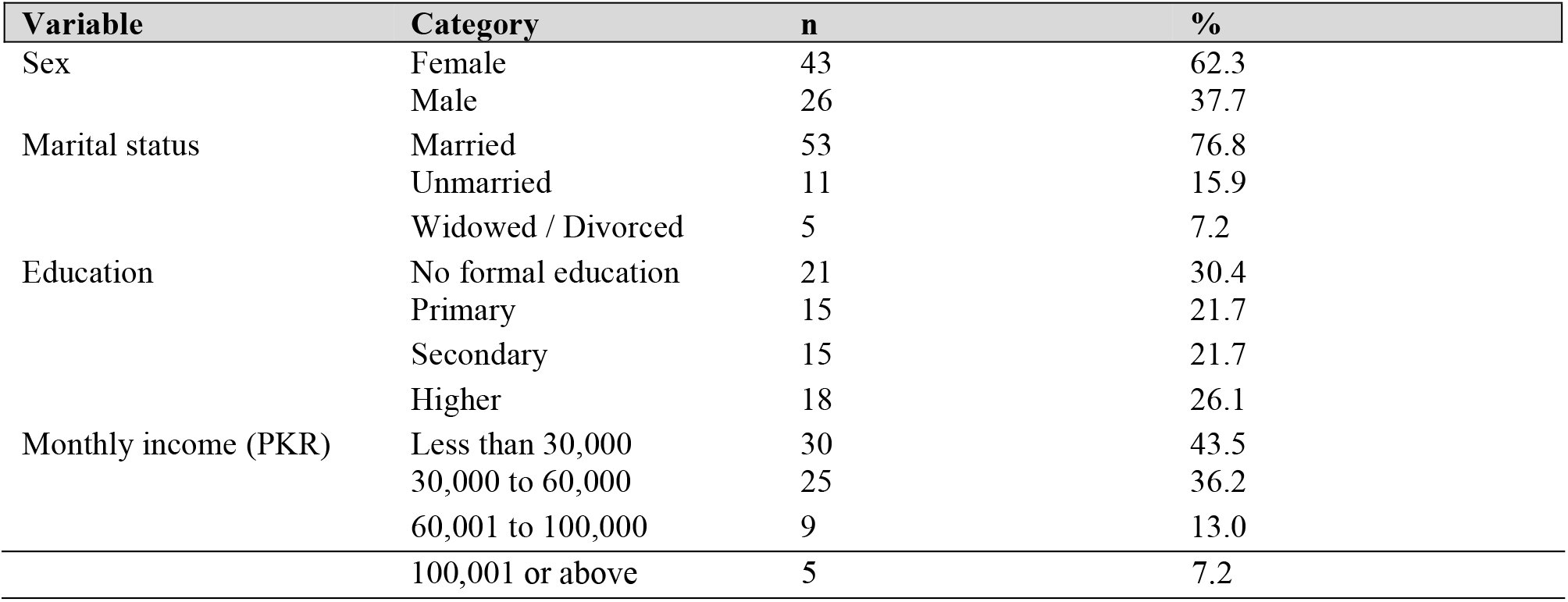
Socio-demographic characteristics of study participants (n = 69)

### Health seeking behaviour

Among 69 participants, 68 (98.6%) consulted an informal provider as their first point of contact. The single participant who sought formal care first was exceptional. The most frequent first providers were unqualified practitioners (35, 50.7%), followed by homeopaths (12, 17.4%) and herbal practitioners (20, 29.0%). The most common reason for choosing informal care was trust in the provider (30, 43.5%), followed by proximity (17, 24.6%), low cost (11, 15.9%), and family recommendation (10, 14.5%). The first provider distribution and selection criteria are shown in Table 2 as follows.

**Table 2.**
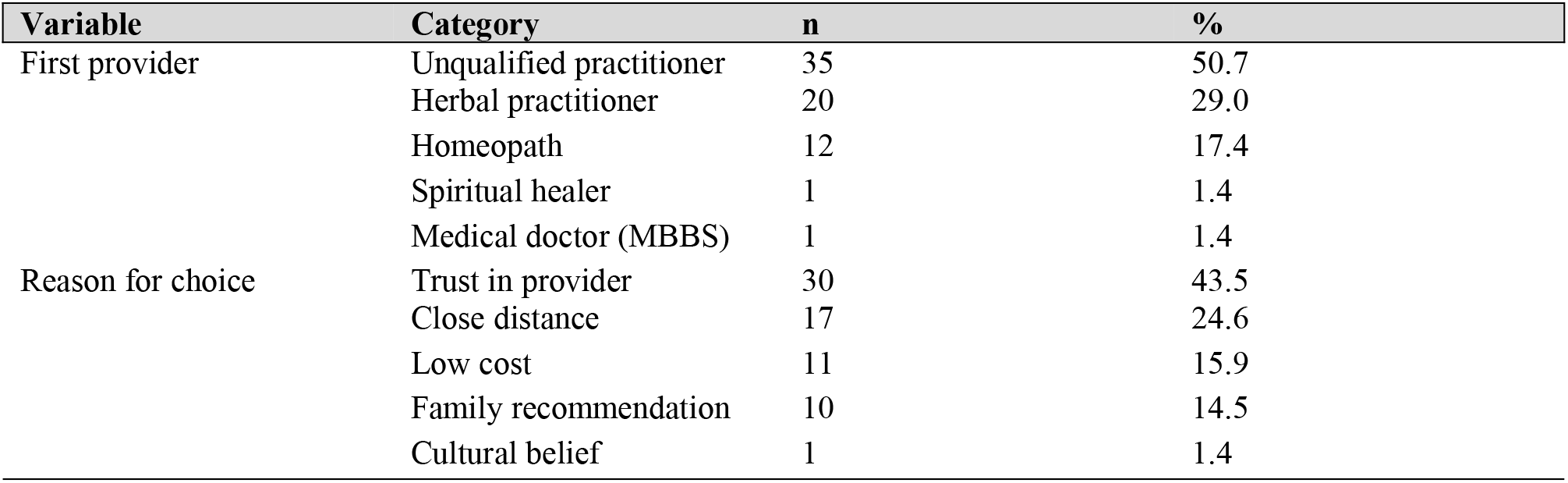
First provider consulted and reasons for choice (n = 69)

| Variable | Category | n | % |
| --- | --- | --- | --- |
| First provider | Unqualified practitioner | 35 | 50.7 |
|  | Herbal practitioner | 20 | 29.0 |
|  | Homeopath | 12 | 17.4 |
|  | Spiritual healer | 1 | 1.4 |
|  | Medical doctor (MBBS) | 1 | 1.4 |
| Reason for choice | Trust in provider | 30 | 43.5 |
|  | Close distance | 17 | 24.6 |
|  | Low cost | 11 | 15.9 |
|  | Family recommendation | 10 | 14.5 |
|  | Cultural belief | 1 | 1.4 |

**Table 3.** Chi-square analysis of associations between socio-demographic variables, provider choice, and outcomes (n = 69)

| Association tested | Chi-square value | Degrees of freedom | p-value |
| --- | --- | --- | --- |
| Education level vs provider type (informal vs formal) | 0.002 | 1 | 0.965 |
| Monthly income vs provider type (informal vs formal) | 0.554 | 1 | 0.457 |
| Sex vs provider type (informal vs formal) | 0.066 | 1 | 0.798 |
| Unqualified practitioner (first provider) vs complications | 0.271 | 1 | 0.603 |
| Unqualified practitioner (first provider) vs later hospital visit | 0.119 | 1 | 0.730 |
| Education level vs complications | 0.155 | 1 | 0.694 |
| Monthly income vs complications | 0.000 | 1 | 1.000 |
| Trust as reason for choice vs later hospital visit | 0.685 | 1 | 0.408 |

### Treatment outcomes

Five participants (7.2%) reported no improvement, eleven (15.9%) reported partial improvement, and fifty-three (76.8%) indicated improvement after receiving informal care. Fifteen individuals (21.7%) reported experiencing complications or their condition worsening. Moreover, the same illness required twenty-seven individuals (39.1%) to visit a formal facility.

### Chi-square analysis of associations

Chi-square tests were used to investigate relationships between clinical outcomes and provider type as well as between socio-demographic factors and provider choice. Table 4 provides a summary of the findings. For every tested link, no statistically significant associations were found at the p < 0.05 criterion. Education level was not associated with provider choice (chi-square 0.002, p = 0.965). Similarly, monthly household income (chi-square 0.554, p = 0.457), and sex (chi-square 0.066, p = 0.798) were not associated with selection of informal providers.

Consultation of an unqualified practitioner was neither associated with complications (chi-square 0.271, p = 0.603) nor with requiring subsequent hospital care (chi-square 0.119, p = 0.730). Education level (chi-square 0.155, p = 0.694) and income (chi-square 0.000, p = 1.000) were also not significantly associated with complications. The reason for provider choice (trust versus other reasons) was not significantly associated with subsequent hospitalisation (chi-square 0.685, p = 0.408).

Overall, the absence of statistically significant associations is likely attributable to the near-universal rate of informal care seeking (98.6%), which constrained variance in the primary outcome variable and limited the statistical power of chi-square analysis in this exploratory sample.

### Qualitative findings

Twelve in-depth interviews and two focus groups were subjected to thematic analysis, which produced eight themes. Quotations from representative participants taken directly from the transcripts are used to support each theme.

### Theme 1: Structural and economic barriers to formal health care

The most frequently mentioned structural barrier in both focus groups and the twelve in-depth interviews was financial expense. Instead of just being inconvenient, participants described formal care as practically unavailable. One participant noted: “Government hospitals create a lot of trouble, so I did not have the power to go there… that is why we have to go to nearby alternatives.” Another stated: “They write expensive medicines and also expensive tests.” (IDI2). A participant in IDI8 explained that cost drove her family to a local unqualified provider: “Because they are expensive, and this hospital is nearby. Whenever we have a problem, we go there.” Beyond cost, bureaucratic barriers such as registration queues and complicated slip-obtaining procedures further added to their burden. Focus group participants collectively described government hospitals as open only during daytime hours, requiring patients to travel long distances at night when emergencies arise, a concern that directly contributed to the near-fatal outcome described in IDI4, where a patient with undetected hypertension received an incorrect injection from an unqualified compounder because no formal facility was accessible at 2 am.

### Theme 2: Proximity and convenience as primary determinants of provider choice

Geographic proximity emerged as a distinct and independent determinant of provider choice. When sickness onset was acute, participants from a variety of IDIs reported selecting providers based on immediate availability rather than preference. IDI3 stated that she “could not walk” and needed emergency care, so she went to a nearby unqualified physician for a high fever. IDI4 described consulting a compounder because it “was nearby” and there was no transport to the district hospital at night. IDI11 noted that the village health facility “only opens during the day” and facilities that open around the clock were too distant for residents to practically reach. Focus group participants reinforced this, with one participant stating: “Distance to hospitals and lengthy tests discourage people from going there.” The pattern among participants indicates that the first point of contact is determined by proximity as a practical restriction rather than a desire, especially in acute sickness, presentations at night, and homes without dependable transportation.

### Theme 3: Trust, familiarity, and social network influence

In contrast to clinical credentials, trust in informal providers was based on long-term interpersonal acquaintance and was expressed consistently among IDIs. Participants described long-standing relationships with local hakeems, unqualified practitioners, and homeopaths as generating a form of relational trust that formal providers had not offered. IDI1 explained: “A woman from my neighborhood told me about that Hakeem. She said that if I took medicine from there, I would feel better.” IDI5 stated: “I have faith in him. I go to him every time because I trust him.” IDI10 described returning repeatedly to a neighbourhood Hakeem because “he sees carefully and listens attentively.” Family guidance was a closely related driver: IDI7 explained that the visit to a homeopath was initially “according to their wish,” and IDI10 described being directed by an elder at home. Focus group discussions revealed that family disputes occasionally occurred when some members supported traditional practitioners while others favoured formal treatment, suggesting that the social negotiation of care seeking involves both intra-household and community-level processes.

### Theme 4: Cultural and religious normalisation of informal practices

The perceptions of many participants about the validity of homeopathic treatment, herbal remedies, and spiritual healing were influenced by cultural and religious perspectives. The use of taweez (protective amulets) and dam (Quranic recitation over water) was reported in both focus group discussions as a culturally normal initial response to illness, especially for diseases thought to have spiritual or supernatural components. FGD2 participants described a Pir nearby whose amulets many families routinely sought, noting that “cultural beliefs strongly influence these choices.” Participants in IDI4 and FGD1 described hakim treatment and herbal treatments as family-passed customs that represent cumulative communal knowledge rather than a break from logical care. One FGD2 participant noted that homeopathic medicines were preferred because they are “affordable, sweet in taste, and do not have side effects,” framing this choice within a broader cultural narrative of naturalness and safety. For participants who used spiritual healers, treatment was experienced not merely as a medical act, but as a source of reassurance and social belonging, consistent with the cultural faith described across both FGD groups.

### Theme 5: Poor doctor-patient communication and interpersonal dissatisfaction in formal settings

Dissatisfaction with the quality of communication in formal health settings was expressed across all twelve in-depth interviews and in both FGDs, making this the most pervasive theme in the qualitative dataset. Participants described hurried consultations in which doctors wrote prescriptions without listening to the full history. IDI1 stated: “Doctors do not listen properly. They just write a prescription quickly and do not pay attention to what the patient is saying.” IDI9 observed that “patients at hospitals are not treated properly. Doctors don’t even listen to patients’ problems.” IDI10 described a pattern of doctors who “just make the prescription quickly and send us somewhere else.” The contrast with informal providers was stark: participants who described positive experiences with hakeems and homeopaths consistently cited attentiveness as the distinguishing quality, as illustrated by IDI10’s praise that the Hakeem “sees carefully and listens attentively,” and IDI11’s description of a provider whose “behaviour was very good; he listens carefully.” The desire for respectful, patient-centred communication emerged as the single most frequently recommended improvement, with participants across almost all IDIs and FGDs independently stating that improved doctor behaviour would be the most impactful change in encouraging formal care use.

### Theme 6: Perceived safety and naturalness of alternative remedies

Several participants across IDIs and FGDs expressed the belief that herbal medicines and homeopathic remedies are inherently safer than allopathic treatments because they are natural and free from side effects. FGD2 participants stated: “Homeopathic medicines are affordable, sweet in taste, and do not have side effects, so people choose them over conventional doctors. Cultural acceptance of homeopathy also plays a role.” This perception was widespread even among participants who held positive views about MBBS doctors, suggesting that the preference for alternative remedies is not simply a consequence of distrust in bio-medicine but an active belief in the superiority of natural treatments for certain conditions. In contrast to traditional remedies, IDI1 characterized allopathic medications as potent and possibly dangerous. This framing was repeated by several FGD1 participants, who stated that local practitioners were preferred for common ailments due to the negative effects of formal medications. This theme illustrates a form of risk perception that operates independently of access constraints and has implications for health communication strategies.

### Theme 7: Low health literacy and awareness deficits about provider qualifications

A prominent theme that emerged from IDI2, IDI5, IDI6, IDI7, IDI8, and IDI11, as well as both FGDs, was that many community members could not distinguish between qualified MBBS doctors and unqualified practitioners who dispensed allopathic medicines. IDI2 explained: “Some people in our area do not even know that an MBBS doctor is someone who is properly trained. They just see the doctor giving medicines or injections and assume that it is fine.” IDI6 noted: “Some people don’t know that local providers are not qualified MBBS doctors. They assume taking medicines will make them better.” IDI5 and IDI7 independently repeated nearly identical observations. Participants unanimously identified awareness campaigns as a high-priority intervention, with IDI6 recommending: “If information is given, people will go to MBBS doctors.” IDI8 suggested that witnessing adverse outcomes from unqualified providers could serve as an effective community-level trigger for behaviour change. This deficit in health literacy about provider qualifications represents a structural vulnerability that allows the informal sector to retain patients who might otherwise prefer formal care if they understood the distinction.

### Theme 8: Treatment-related harm and delayed escalation to formal care

Several participants described clinical harm directly attributable to informal care, and a pattern of delayed escalation to formal services despite recognising treatment failure. IDI3 described developing an allergy and worsening hypertension following medicines from an unqualified provider, yet did not seek formal care until her internal problems kept increasing over an extended period of time. IDI4 provided the most severe case: a compounder administered treatment for presumed hypotension in a patient who actually had hypertension, resulting in a cardiac event requiring emergency transfer to DHQ Faisalabad. IDI9 developed loose stools following herbal treatment for uric acid, but only sought formal care once these secondary symptoms became intolerable. IDI12 described partial improvement followed by side effects and eventual worsening. These testimonies show a pattern where patients return to the same informal provider after initial problems before seeking qualified care, and where the barrier to ascending to formal care is defined by deterioration rather than lack of initial improvement. The qualitative data thus extend the quantitative finding that 21.7% of participants experienced complications by revealing that harm is often experienced as a graduated process rather than an acute event, with multiple opportunities for intervention that the current system does not capture.

## DISCUSSION

This study demonstrates that nearly all adults in peri-urban and rural Faisalabad pursue informal health care. Since 98.6% of participants initially accessed guidance from an informal provider, the data demonstrate that informal providers have a deeply embedded functional role in these communities. By filling gaps caused by cost, physical unreachability, bureaucratic barriers, and deficient interpersonal care, this care becomes a preferred choice. These tendencies mediate structural and cultural problems that require wilful governmental attention and cannot be explained by a lack of awareness of formal services. This study’s convergent mixed-methods design produced a more comprehensive depiction of health-seeking behavior by allowing qualitative narratives to validate the quantitative trends.

The national data showing abundant quackery in Pakistan’s low-income populations is concordant with the prevalence of unqualified practitioners as initial contact (50.7%)[5]. According to Andersen’s Behavioural Model, the high level of trust placed in informal providers (43.5% of participants) reflects community credibility based on neighbourhood familiarity and word-of-mouth recommendation [10]. In this situation, trust functioned as a highly social and relational construct that resulted from long-term community ties. As a result, if departments do not properly address the relational aspect of provider trust, programs that offer accurate data about capabilities may only partially act on behaviour.

Chi-square analysis did not identify statistically significant associations between any of the tested socio-demographic variables and provider choice, complications, or subsequent hospitalization (Table 4). This finding is analytically important rather than merely negative. When 98.6% of participants consult informal providers first, there is insufficient variance in the outcome to detect associations with any predictor variable in a sample of this size. The result confirms that informal care seeking is not concentrated among any particular demographic subgroup defined by sex, education, or income but is a near-universal community norm cutting across these characteristics. Programmes targeting only low-income or low-education groups would therefore miss the broad social norm driving informal care use. A larger study with greater variance in formal care use, powered to detect associations by regression modelling, is needed. Thematic analysis of all twelve in-depth interviews and both focus group discussions identified eight themes, substantially expanding the analytical depth of the qualitative component and revealing dimensions not captured in smaller qualitative samples.

Even though 21.7% of participants reported problems, the Health Belief Model provides additional explanatory value for understanding perseverance with informal care [9]. The stated intensity of illness was consistently overruled by perceived barriers to gaining access to formal healthcare, including cost, bureaucratic difficulties, and negative prior experiences with medical facilities, for most participants. As a result, 39.1% of the sample population delayed seeking official care until circumstances worsened. This is in line with studies conducted in Pakistan on cancer patients, which found that early reliance on traditional medicine was associated with a poorer prognosis and delayed presentation [4], as well as studies from Karachi’s mental health care channels, where patients alternated between spiritual and medical professionals before receiving the proper treatment [6].

79.7% of participants reported household earnings below PKR 60,000 per month. Pakistan has some of the highest out-of-pocket medical costs in the area, and low-income families often cannot pay the consultation fees, diagnostic tests, and medications administered at formal institutions during a single disease episode. Policy responses that solely address the supply side of formal health care, such as building new facilities, will have minimal impact unless affordability is concurrently addressed through subsidized medications, waived primary care consultation fees, and protected budgets for necessary medications in public facilities.

The cultural dimension of informal care seeking documented here aligns with findings from Sindh, where shrine-based healing was normalized within community frameworks for illness [3]. Health communication strategies must engage respectfully with traditional frameworks. Rather than relying solely on facility-based health education, community health workers with a reputation within local networks may be in a better position to close the gap between conventional wisdom and evidence-based treatment. Without portraying early formal care seeking as a rejection of faith or tradition, religious and community leaders could be involved as collaborators in spreading the message.

The findings suggested that participants did not simply lack awareness of formal services; many had previously used them and found the experience interpersonally discouraging. Factors that decreased willingness to return included hurried consultations, inattentive attention, and contemptuous attitudes. This is consistent with wider literature indicating that patient-centred communication is a key determinant of health-care utilisation and treatment adherence in low-resource settings. Our study recommends that in Pakistan, community health engagement, collaborative decision-making, and communication skills should all be systematically taught in undergraduate medical education. These competencies should also be given priority in continuing professional development for physicians who practice in public institutions.

Social desirability bias may have influenced the reporting of complications and treatment outcomes. The one-month data collection period does not capture seasonal variation in illness burden or health seeking. The qualitative sample was modest, even though it reached preliminary topic saturation. Larger probability samples, longitudinal follow-up to record health outcomes after informal care, and quantitative analysis of relationships between socio-demographic characteristics and provider choice should all be used in future studies. Research comparing Punjab’s rural and urban areas would shed further light on the function of geographic access as a separate factor influencing the use of informal care.

## CONCLUSIONS

In peri-urban and rural Faisalabad, seeking health care informally is almost universal and is powered by a combination of structural barriers, cultural beliefs, poor doctor-patient interactions, and economic challenges. For most of this population, homeopaths, herbalists, and unqualified practitioners serve as the de facto primary care system. The significance of addressing these pathways for public health is stressed by the interconnected burden of complications and subsequent hospitalization. Multi-pronged interventions that concurrently address primary care quality, affordability, geographic access, and cultural sensitivity are needed. Future doctors should be prepared by medical education to work successfully in these community settings.

## ARTICLE SUMMARY

### Strengths and limitations

A convergent parallel mixed-methods design was used, integrating a structured community survey with in-depth interviews and focus group discussions, which strengthens the credibility of findings through methodological triangulation.

The study was conducted in a real community setting in peri-urban and rural Faisalabad, capturing health-seeking behaviour in a context that is underrepresented in existing mixed-methods research on informal care in Punjab.

The use of the validated I-CAM-Q instrument and theoretically grounded qualitative guides based on the Health Belief Model and Andersen’s Behavioural Model ensured systematic and replicable data collection.

Purposive sampling and a relatively small sample of 69 survey participants limit the generalisability of findings to wider populations or other regions of Punjab.

The single-month data collection period and reliance on self-reported outcomes may not fully capture seasonal variation in health seeking or be free from social desirability bias.

The very high proportion of participants reporting informal healthcare use resulted in an imbalanced distribution of the primary outcome variable, which limited statistical power and constrained chi-square analyses.

## Data Availability

All data produced in the present study are available upon reasonable request to the authors

## DECLARATIONS

### Ethics approval and consent to participate

Approval was obtained from the IRB of King Edward Medical University, Lahore. Written informed consent was obtained from all participants.

### Authors’ contributions

SH designed the study, collected and analysed data, and drafted the manuscript. MM contributed to data collection and qualitative analysis. S conceptualised and supervised the study and critically revised the manuscript. All authors approved the final version.

## Acknowledgements

The authors thank the community members of Makkuana and surrounding villages of Faisalabad for their participation.

